# Hiatal Hernia Size and De Novo Gastroesophageal Reflux Disease After Sleeve Gastrectomy: A Single-Center Retrospective Study

**DOI:** 10.64898/2026.08.31.26361833

**Authors:** Eros Rafael Ricarte Almeida, Carlos Javier Mata Quíntero, Jaime Sesma Cházaro, Cristina Peralta Rivera, Camila Dennys Arteaga González

**Author notes:** **Corresponding author:** Eros Rafael Ricarte Almeida, Hospital Central Norte PEMEX, Mexico City, Mexico.

## Abstract

**Background:** Sleeve gastrectomy is the most frequently performed bariatric procedure worldwide but is associated with the development of de novo gastroesophageal reflux disease (GERD). Hiatal hernia has been identified as a relevant anatomical factor in postoperative reflux, although most studies evaluate it dichotomously without analyzing whether its size influences GERD risk. The aim was to evaluate the association between preoperative hiatal hernia size and de novo GERD after sleeve gastrectomy.

**Methods:** Retrospective, single-center, observational study of patients undergoing sleeve gastrectomy at Hospital Central Norte de Petróleos Mexicanos (2018–2025). Demographic and clinical characteristics, endoscopic classification of hiatal hernia size (small <2 cm, medium 2.1–4 cm, large >4 cm), and evidence of de novo GERD were analyzed using descriptive statistics, Fisher’s exact test, odds ratio (OR) estimation with 95% confidence intervals (CI), and binary logistic regression. Statistical significance was set at p<0.05.

**Results:** Fifty-six patients were included (mean age 48.3 ± 8.1 years; 67.9% male). Hiatal hernia classification was conclusive in 46 patients (82.1%): 63.0% no hernia, 4.3% small, 30.4% medium, and 2.2% large. De novo GERD occurred in 14.0% of patients without preexisting GERD (6/43). No significant association was found between hiatal hernia size and de novo GERD (Fisher p=0.515). In the reduced logistic model, neither hiatal hernia (medium/large vs. absent/small; OR 3.47; 95% CI 0.50– 29.43; p=0.207) nor age (OR 1.02; 95% CI 0.90–1.13; p=0.754) was significantly associated. No evaluated factor (sex, smoking, alcohol, age) reached significance.

**Conclusions:** In this cohort, no statistically significant association was demonstrated between preoperative hiatal hernia size and de novo GERD after sleeve gastrectomy; however, the low number of events limits the ability to exclude a clinically relevant association. These findings are compatible with a multifactorial mechanism rather than with the isolated presence of this finding. Prospective studies with larger sample sizes and standardized reflux assessment instruments are required to confirm these results.

## Introduction

Obesity is a highly prevalent chronic disease worldwide, and bariatric surgery has become the most effective treatment for achieving sustained weight loss and the improvement or remission of associated comorbidities. Among the available techniques, sleeve gastrectomy has become the most frequently performed bariatric procedure worldwide, owing to its lower technical complexity, good safety profile, and favorable weight-loss outcomes [1].

Despite these advantages, one of the main drawbacks of sleeve gastrectomy is its relationship with gastroesophageal reflux disease (GERD). Conversion of the stomach into a narrow tubular reservoir modifies the anatomy of the esophagogastric junction, alters gastric distensibility, and may compromise the competence of the lower esophageal sphincter, favoring the appearance of de novo reflux or the worsening of preexisting reflux [2,3,4]. Among the anatomical factors involved, hiatal hernia has been consistently identified in the literature as a relevant element in the pathophysiology of postoperative reflux [4,5,6].

However, most available evidence analyzes hiatal hernia dichotomously (present/absent), without exploring whether its size modifies the risk of development or the severity of GERD. This knowledge gap limits clinical decision-making in the preoperative period, particularly regarding the indication for concomitant hiatal repair or the selection of the most appropriate bariatric procedure for each patient [4,5,6,7].

At Hospital Central Norte de Petróleos Mexicanos (PEMEX), where bariatric surgery is performed routinely, no local evidence on this association was previously available. Therefore, the present study aimed to analyze the relationship between preoperative hiatal hernia size and evidence of de novo GERD in patients undergoing sleeve gastrectomy, in order to provide information that could improve patient selection and surgical decision-making in this setting.

## Materials and methods

### Study design and population

A retrospective, single-center, observational study was conducted in patients who underwent sleeve gastrectomy at Hospital Central Norte de Petróleos Mexicanos between 2018 and 2025. Information was obtained through review of electronic and paper medical records, including operative notes, preoperative endoscopy (panendoscopy) reports, outpatient follow-up notes, and pharmacy records, and was captured in an electronic database designed specifically for this study. Postoperative follow-up was standardized at 12 months, corresponding to the annual reassessment contemplated in the institutional bariatric surveillance protocol; this window was used as the reference for determining de novo GERD and PPI use.

### Inclusion and exclusion criteria

We included patients who underwent sleeve gastrectomy at the institution during the study period; older than 18 years; with a complete medical record; with a documented endoscopic evaluation; with a record of the presence and size of hiatal hernia; and with postoperative follow-up including clinical and/or endoscopic data allowing identification of evidence of GERD. We excluded patients who underwent a bariatric procedure other than sleeve gastrectomy and those with incomplete records. Preexisting GERD and esophagitis were not exclusion criteria for the cohort; their presence was handled analytically through the definition of the analysis sets (see Statistical analysis).

### Variables

The primary outcome was de novo GERD, defined (in patients without preoperative reflux) as the appearance, documented in the medical record, of at least one of the following: new-onset typical reflux symptoms, a clinical diagnosis of GERD, compatible endoscopic findings (e.g., esophagitis), and/or initiation of PPI treatment for a symptomatic indication. Prophylactic or empirical postoperative PPI use, prescribed per protocol and not for symptomatic reflux, was not considered by itself a criterion for de novo GERD. Given the recognized heterogeneity in the operational definition of GERD across studies, the concept of “evidence of GERD” was used, integrating documented clinical and endoscopic findings rather than relying on a single diagnostic modality [4,6,8].

Hiatal hernia size was classified by endoscopy as small (<2 cm), medium (2.1–4 cm), or large (>4 cm). Other variables included age, sex, initial weight, excess weight, percentage of excess weight loss at one year, psychiatric history, postoperative use of proton pump inhibitors (PPIs), smoking, and alcohol consumption.

### Statistical analysis

Continuous variables were summarized as mean ± standard deviation or median and interquartile range (IQR) according to their distribution; categorical variables were expressed as frequencies and percentages. The association between hiatal hernia size and de novo GERD was evaluated with the chi-square test or Fisher’s exact test as appropriate, estimating the odds ratio (OR) with a 95% confidence interval (CI). A bivariate analysis and a binary logistic regression model were performed to identify factors independently associated with de novo GERD. Analyses were structured in three nested sets: (I) the full cohort (n=56), used to describe baseline characteristics; (II) the de novo GERD set, comprising patients without preoperative GERD (n=43), used to estimate the frequency of new-onset reflux; and (III) the primary analysis set, comprising patients without preoperative GERD and with an endoscopically evaluable hiatal hernia (n=34), used to evaluate the association between hiatal hernia size and de novo GERD. Statistical significance was set at p<0.05. Analyses were performed in R/RStudio.

### Ethical considerations

The study was conducted in accordance with the Declaration of Helsinki and Article 17 of the Regulations of the General Health Law on Health Research (Mexico), and was classified as risk-free research. The protocol was approved by the institutional Research Committee (COFEPRIS registration 18CL09002035), which granted a waiver of individual informed consent given the retrospective design and the use of an anonymized database.

## Results

Fifty-six patients who underwent sleeve gastrectomy between 2018 and 2025 were included. The mean age was 48.3 ± 8.1 years (median 46.5; IQR 43.0–54.0). All patients completed a standardized 12-month postoperative follow-up, in accordance with the institutional bariatric surveillance protocol. Thirty-eight patients (67.9%) were male and 18 (32.1%) female. The mean initial weight was 116.7 ± 22.4 kg, with a mean excess weight of 58.9 ± 21.0 kg. Twenty-two patients (39.3%) achieved excess weight loss greater than 50% at one year, whereas 34 (60.7%) had less than 50%. Psychiatric history was documented in 8 patients (14.3%), smoking in 21 (37.5%), and alcohol consumption in 10 (17.9%) (Table 1).

**Table 1.**
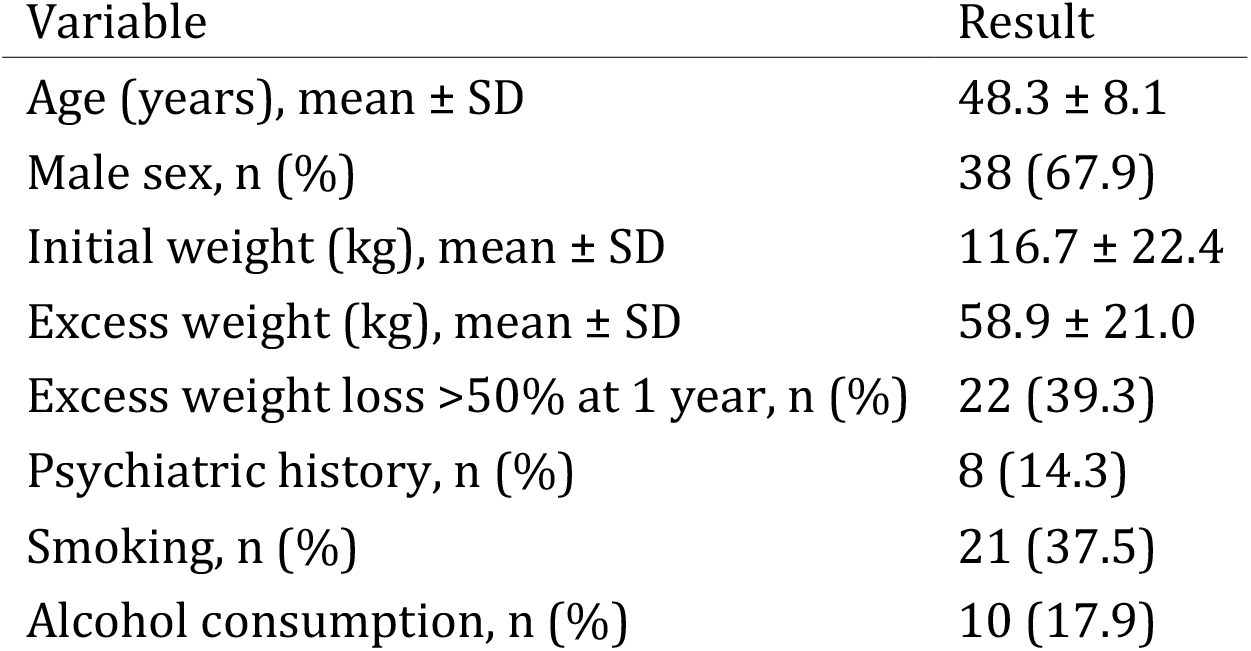
Demographic and clinical characteristics of the study population (n=56)

| Variable | Result |
| --- | --- |
| Age (years), mean ± SD | 48.3 ± 8.1 |
| Male sex, n (%) | 38 (67.9) |
| Initial weight (kg), mean ± SD | 116.7 ± 22.4 |
| Excess weight (kg), mean ± SD | 58.9 ± 21.0 |
| Excess weight loss >50% at 1 year, n (%) | 22 (39.3) |
| Psychiatric history, n (%) | 8 (14.3) |
| Smoking, n (%) | 21 (37.5) |
| Alcohol consumption, n (%) | 10 (17.9) |

Endoscopic evaluation of hiatal hernia was conclusive in 46 of the 56 patients (82.1%); the remaining 10 were excluded from hiatal hernia–related analyses because they lacked an evaluable endoscopic study or had an unclassifiable finding. Of the 46 evaluable patients, 29 (63.0%) had no hiatal hernia, 2 (4.3%) had a small hernia (<2 cm), 14 (30.4%) a medium hernia (2.1–4 cm), and 1 (2.2%) a large hernia (>4 cm) (Table 2).

**Table 2.** Hiatal hernia classification (n=46 evaluable)

| Classification | n | % |
| --- | --- | --- |
| Absent | 29 | 63.0 |
| Small (<2 cm) | 2 | 4.3 |
| Medium (2.1–4 cm) | 14 | 30.4 |
| Large (>4 cm) | 1 | 2.2 |

In the de novo GERD set (the 43 patients without preexisting GERD, in whom the appearance of de novo reflux can be properly evaluated), the frequency of de novo GERD was 14.0% (6/43), with PPI use in 65.1% (28/43). In the full cohort (n=56), 64.3% received PPI treatment at some point during follow-up (Figure 1).

**Figure 1.**
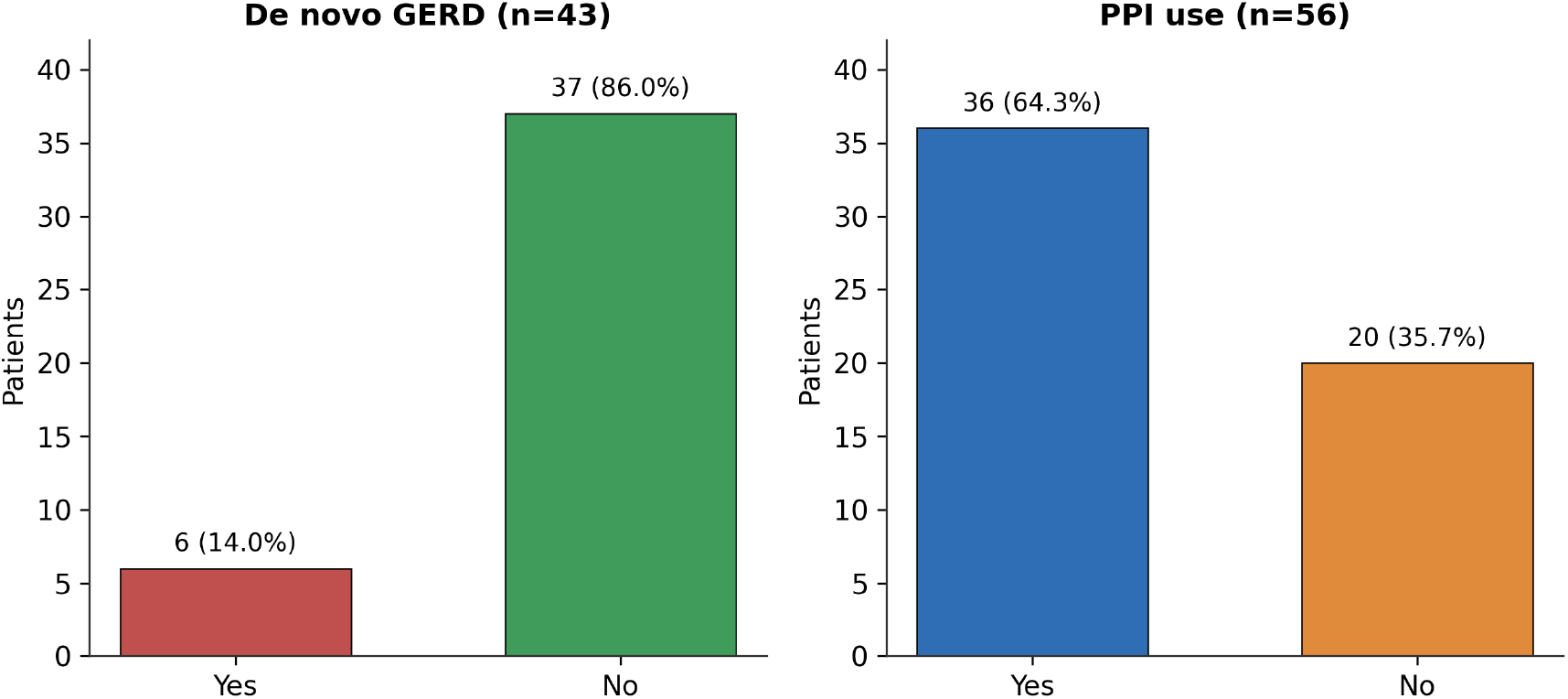
De novo GERD and proton pump inhibitor (PPI) use among patients undergoing sleeve gastrectomy. Left panel: proportion of patients with de novo GERD in the de novo GERD set (n=43). Right panel: proportion of patients with PPI use in the full cohort (n=56). GERD: gastroesophageal reflux disease; PPI: proton pump inhibitor.

When analyzing the association between hiatal hernia classification and the presence of de novo GERD in the 46 patients with an evaluable hiatal hernia, no statistically significant association was found (p=0.515). When grouping the categories into “absent/small” versus “medium/large” to estimate an odds ratio (reference category: absent/small), no statistical significance was observed either. By category, the percentage of patients with de novo GERD was 6.9% in absent (n=29), 0.0% in small (n=2), 21.4% in medium (n=14), and 0.0% in large (n=1) (Figure 2). The primary analysis, restricted to patients without preexisting GERD and with an evaluable hiatal hernia (n=34), was consistent (Fisher’s exact test p=0.377). In both scenarios the confidence intervals were extremely wide, reflecting the low number of observed events (2–3 cases of de novo GERD in the medium/large hernia categories), which limits statistical power.

**Figure 2.**
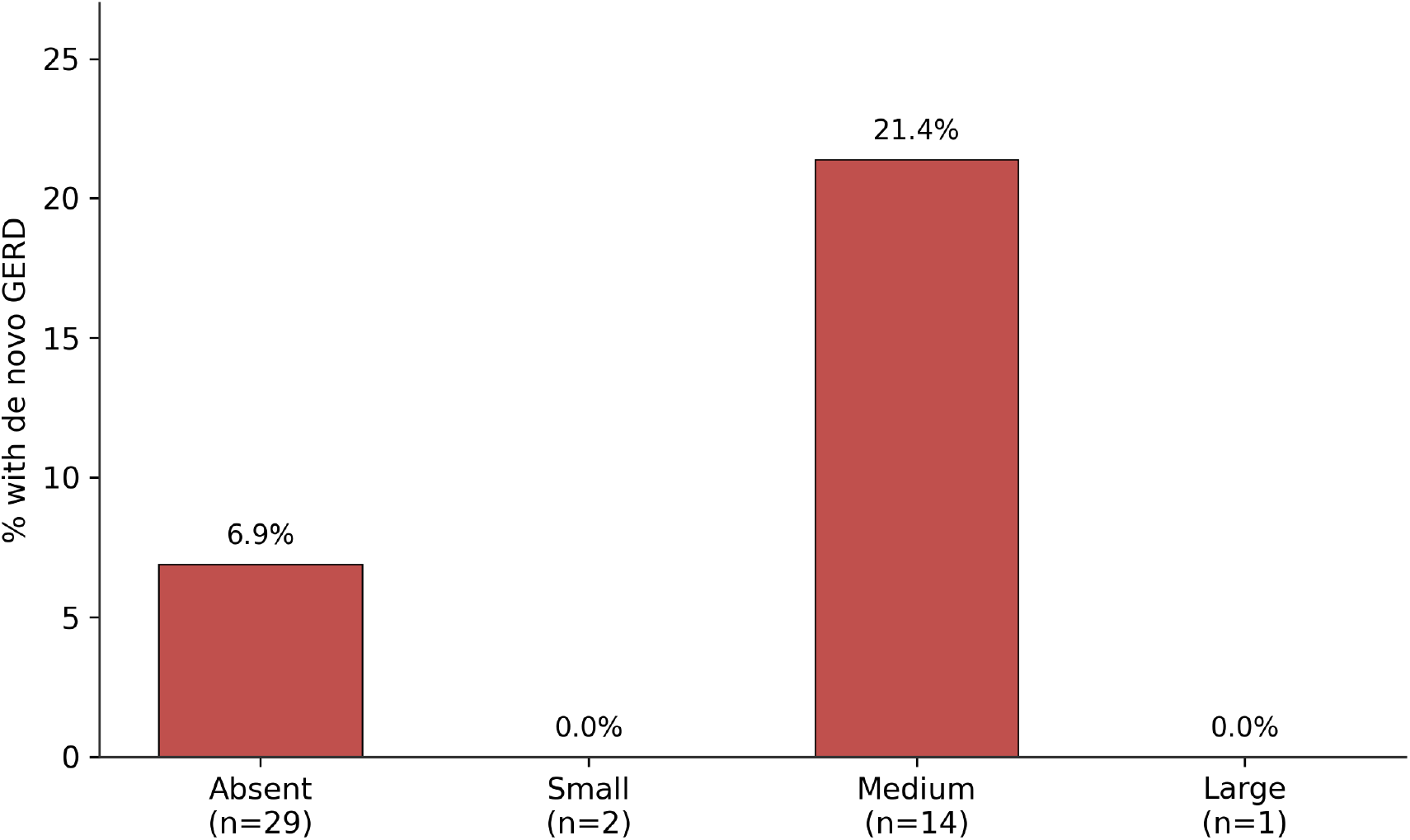
Percentage of patients with de novo GERD according to hiatal hernia size classification (n=46 evaluable). GERD: gastroesophageal reflux disease.

In the bivariate analysis, none of the evaluated variables showed a statistically significant association with de novo GERD: male sex (OR 0.59; 95% CI 0.12–2.96; p=0.52), smoking (OR 0.40; 95% CI 0.08–1.99; p=0.262), alcohol consumption (OR 1.35; 95% CI 0.14–12.64; p=0.793), or age as a continuous variable (OR 1.04 per year; p=0.449) (Figure 3).

**Figure 3.**
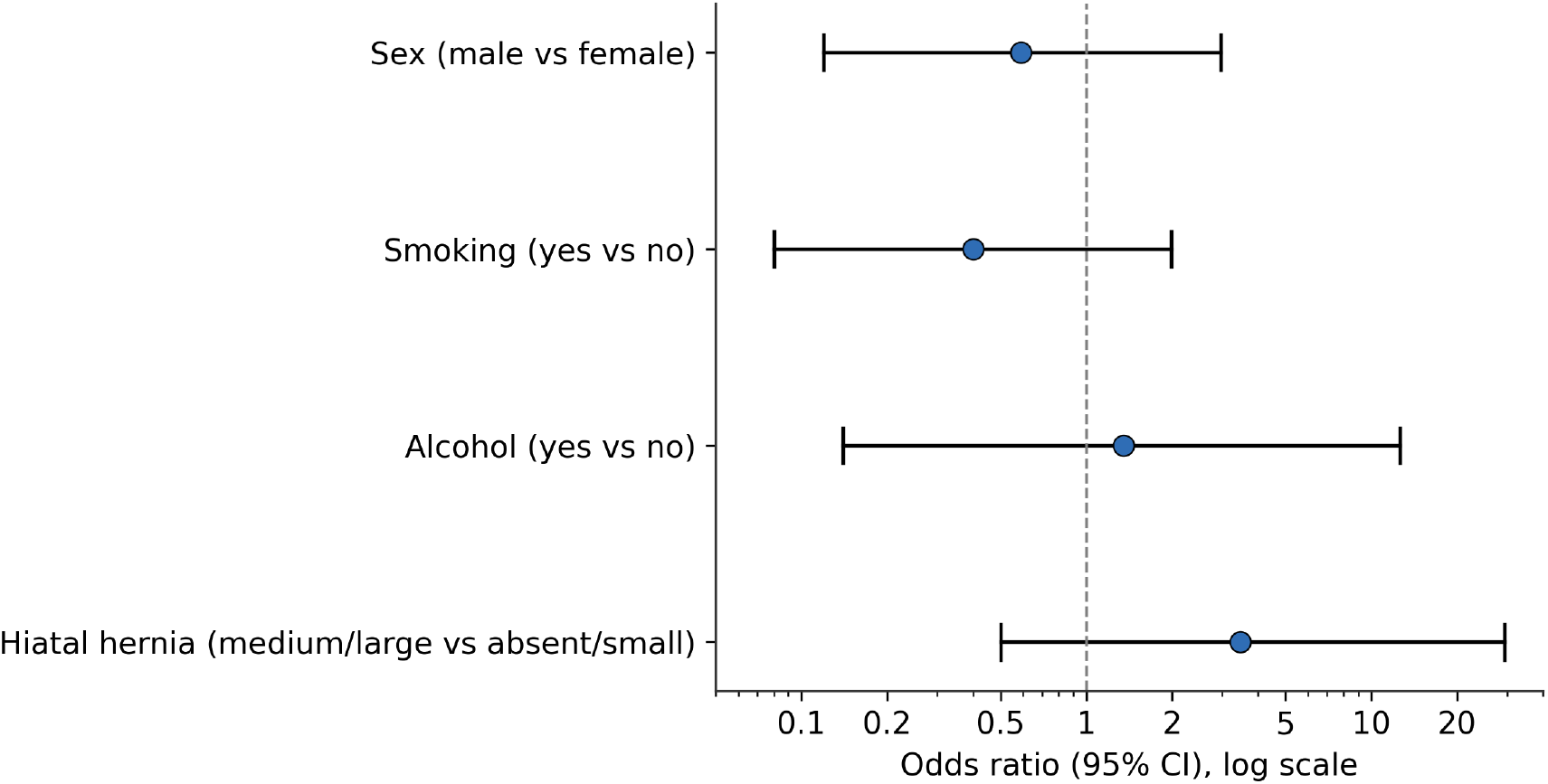
Odds ratios (OR) and 95% confidence intervals for factors associated with de novo GERD. Sex, smoking, and alcohol correspond to crude bivariate estimates; the hiatal hernia estimate corresponds to the reduced logistic regression model. The dashed line indicates OR = 1.

For the multivariate analysis, a binary logistic regression model was initially evaluated. However, owing to the low frequency of de novo GERD (6 events) and the presence of quasi-complete separation in the psychiatric comorbidity variable (0/8), a reduced exploratory model including age and hiatal hernia was fitted. In this analysis, neither hiatal hernia (medium/large vs. absent/small [reference]; OR 3.47; 95% CI 0.50–29.43; p=0.207) nor age (OR 1.02; 95% CI 0.90–1.13; p=0.754) showed a statistically significant association with de novo GERD.

## Discussion

In this retrospective study of 56 patients undergoing sleeve gastrectomy, the frequency of de novo GERD was 14.0% (6/43) in patients without preexisting GERD, and no statistically significant association was found between preoperative hiatal hernia size and the development of de novo GERD, either in the bivariate analysis (p=0.515) or in the exploratory logistic regression model (medium/large vs. absent/small hernia [reference]; OR 3.47; 95% CI 0.50–29.43; p=0.207). However, the low number of events (6 cases) limits the ability to exclude a clinically relevant association.

Our findings are consistent with those reported by a multiethnic Asian cohort study, which also found no significant association between the presence of a small hiatal hernia and the development of de novo GERD after vertical sleeve gastrectomy (RR 0.682; 95% CI 0.419–1.111; p=0.125); similarly to our data, the frequency of de novo GERD was even lower in the group with hiatal hernia (57.1%) than in the group without it (76.4%) [9]. That same group concluded that the presence of a small hiatal hernia should not be a reason for exclusion from vertical sleeve gastrectomy, nor does it by itself justify concomitant hiatal repair.

However, other studies report opposite results. One of them, which surgically evaluated the diaphragmatic crura after sleeve gastrectomy, found that the presence of hiatal hernia was indeed associated with a higher frequency of GERD (56.8% vs 21.4%) and PPI use, with a statistically significant difference [10]. Similarly, a prospective study of 217 patients found that mild preoperative esophagitis and small hiatal hernia significantly increased the probability of postoperative GERD [6]. This discrepancy between studies suggests that the relationship between hiatal hernia and de novo GERD after sleeve gastrectomy is not fully elucidated and probably depends on methodological factors such as the timing and method of hernia evaluation (preoperative endoscopy vs. intraoperative finding), the follow-up time, and the operational definition of GERD used in each study.

It is also noteworthy that the frequency of de novo GERD observed in our population (14.0%) was considerably lower than that reported in much of the international literature, where figures range between 45% and 76% depending on the study and follow-up time. This difference could be explained by the retrospective nature of our study, which probably underestimates the true incidence of subclinical reflux or reflux not spontaneously reported by the patient. Complementarily, although 64.3% of patients received PPIs at some point during follow-up, only 14.0% met criteria for de novo GERD; this discrepancy probably reflects the prophylactic or empirical use of PPIs after sleeve gastrectomy (often prescribed per postoperative protocol rather than for symptomatic reflux), so PPI use overestimates the true frequency of GERD and was not used as the sole diagnostic criterion.

From a pathophysiological standpoint, the development of GERD after sleeve gastrectomy has been explained as a multifactorial phenomenon: conversion of the stomach into a narrow tube reduces its distensibility and increases intragastric pressure, while dissection of the angle of His and the esophagogastric junction during resection of the greater curvature may weaken the antireflux barrier regardless of whether an anatomical hiatal hernia is present [4,11]. This could explain why, in our series and others, hiatal hernia size alone does not consistently predict which patients will develop de novo GERD; postoperative reflux appears to depend more on the global mechanical alteration of the esophagogastric junction than on the isolated finding of hiatal hernia on preoperative endoscopy.

### Limitations

This study has several limitations. First, the retrospective, single-center design limits data quality: in 17.9% of patients (10/56) it was not possible to classify the hiatal hernia due to the lack of an evaluable endoscopic study or an unclassifiable finding, which reduced the useful sample size. Second, the number of de novo GERD events (6 cases) was very low, which limited statistical power to detect associations of modest magnitude and required restricting the logistic regression model to an exploratory two-variable analysis, given the quasi-complete separation of data with the psychiatric comorbidity variable. Third, the absence of a standardized, prospective instrument for the diagnosis of GERD (relying instead on retrospective clinical records) may have underestimated the true frequency of the outcome. Finally, because follow-up was standardized at 12 months, cases of later-onset de novo GERD (after the first postoperative year) may not have been captured.

Despite these limitations, our findings provide additional evidence, consistent with part of the existing literature, that hiatal hernia size identified on preoperative endoscopy should not be considered, by itself, a reliable predictor of de novo GERD after sleeve gastrectomy, nor a single criterion for deciding concomitant hiatal repair or the choice of an alternative bariatric procedure. Prospective, multicenter studies with larger sample sizes are required, incorporating standardized reflux assessment instruments (validated questionnaires, 24-hour pH monitoring, or high-resolution esophageal manometry) and systematic medium- and long-term follow-up, to more robustly clarify the true role of hiatal hernia and other mechanical factors in the development of de novo GERD after this procedure.

## Conclusion

In this cohort of 56 patients undergoing sleeve gastrectomy at Hospital Central Norte de PEMEX, the frequency of de novo GERD was 14.0% (6/43) in the subgroup without preexisting GERD, lower than that reported in much of the international literature. No statistically significant association was demonstrated between preoperative hiatal hernia size and de novo GERD in any of the analyses performed, and no other evaluated clinical factor (age, sex, smoking, alcohol) showed a significant association; however, the low number of events limits the ability to exclude a clinically relevant association and substantially reduced statistical power. Taken together, these findings suggest that, in the studied population, hiatal hernia size identified on preoperative endoscopy does not constitute, by itself, a reliable predictor of de novo GERD after sleeve gastrectomy, which is consistent with a multifactorial mechanism related to the global alteration of the antireflux anatomy rather than with the isolated presence of hiatal hernia. Prospective studies with larger sample sizes and standardized reflux assessment instruments are required to confirm these results.

## Declarations

### Author contributions

E.R.R.A. participated in the conception and design of the study, data collection, analysis and interpretation of results, and drafting of the manuscript. C.J.M.Q. participated in the conception of the study, supervision, project administration, and critical revision of the manuscript. J.S.C. contributed to the analysis and interpretation of the data, preparation of the figures, and critical revision of the manuscript. C.P.R. contributed to data collection and critical revision of the manuscript. C.D.A.G. contributed to data collection and critical revision of the manuscript. All authors approved the final version and agree to be accountable for all aspects of the work.

### Data availability

The data supporting the findings of this study are not publicly available because they contain confidential patient clinical information; they may be requested from the corresponding author on reasonable request and will be subject to the corresponding institutional authorizations.

### Ethics approval and consent

The protocol was evaluated and approved by the institutional Research Committee (COFEPRIS registration 18CL09002035), which granted a waiver of individual informed consent given the retrospective design and the use of an anonymized database. The study was conducted in accordance with the Declaration of Helsinki and applicable Mexican regulations (NOM-004-SSA3-2012 and NOM-012-SSA3-2012), and was classified as risk-free research.

## References

1. Angrisani L, Santonicola A, Iovino P, Vitiello A, Higa K, Himpens J, et al. Bariatric surgery worldwide 2018. Obes Surg. 2021;31:192–201.

2. Yeung KTD, Penney N, Ashrafian H, Darzi A, Athanasiou T. Does sleeve gastrectomy expose the distal esophagus to severe reflux?Ann Surg. 2020;271:257–265.

3. Serra FE, Cohen RV. Gastroesophageal reflux disease after sleeve gastrectomy. Dig Med Res. 2024;7:5.

4. Shaker A, Soffer E. Gastroesophageal reflux disease related to laparoscopic sleeve gastrectomy. Mini Invasive Surg. 2025;9. doi:10.20517/2574-1225.2024.105.

5. Veziant J, Benhalima S, Piessen G, Slim K. Obesity, sleeve gastrectomy and gastro-esophageal reflux disease. J Visc Surg. 2023;160(5S):S3–S24.

6. Talebloo J, Gadde KM, Mittal RK, Nguyen NT. Association of GERD with sleeve gastrectomy: An unintended consequence. Curr Diab Rep. 2026;26:3.

7. Mahawar KK, Omar I, Singhal R, Aggarwal S, Allouch MI, Alsabah SK, et al. The first modified Delphi consensus statement on sleeve gastrectomy. Surg Endosc. 2021;35:7027–7033.

8. Memon MA, Osland E, Yunus RM, Hoque Z, Alam K, Khan S. The effect of laparoscopic vertical sleeve gastrectomy and laparoscopic roux-en-Y gastric bypass on gastroesophageal reflux disease: an updated meta-analysis and systematic review of 5-year post-operative data from randomized controlled trials. Surg Endosc. 2024;38:6254–6269.

9. Lye TJY, Ng KR, Tan AWE, Syn N, Woo SM, Lim EKW, Eng AKH, Chan WH, Tan JTH, Lim CH, Yeh CC. Small hiatal hernia and postprandial reflux after vertical sleeve gastrectomy: a multiethnic Asian cohort. PLoS One. 2020;15(11):e0241847.

10. Saba J, Bravo M, Rivas E, Fernández R, Pérez-Castilla A, Zajjur J. Incidence of de Novo Hiatal Hernia after Laparoscopic Sleeve Gastrectomy. Obes Surg. 2020;30(10):3730–3734.

11. Memon MA, Yunus RM, Alam K, Hoque Z, Khan S. Impact of laparoscopic sleeve gastrectomy on lower esophageal sphincter pressure (LESP), lower esophageal sphincter length (LESL) and gastroesophageal reflux disease (GERD) using esophageal function tests (EFTs): a systematic review and meta-analysis. Int J Obes (Lond). 2026;50:127–139.

